# Allele-specific digital PCR enhances precision and sensitivity in the detection and quantification of copy number alterations in heterogeneous DNA samples: an *in silico* and *in vitro* validation study

**DOI:** 10.1101/2023.10.31.23297362

**Authors:** Rogier J. Nell, Mieke Versluis, Willem H. Zoutman, Gregorius P.M. Luyten, Martine J. Jager, Pieter A. van der Velden

## Abstract

The analysis of genetic variation is of crucial importance in cancer care. Measuring copy number alterations, however, remains challenging in heterogenous DNA samples, such as (liquid) biopsies. Using digital PCR, these alterations are classically studied by comparing the abundances of the target of interest to a stable genomic reference. Alternatively, copy numbers may be quantified based on the allelic (im)balance of a heterozygous common single-nucleotide polymorphism (SNP). In this study, the accuracy, practicability, precision and sensitivity of both approaches are evaluated *in silico* using a newly introduced R library ‘digitalPCRsimulations’, and *in vitro* by analysing control samples and uveal melanoma specimens in duplex and multiplex experimental setups.

Our analyses show that both methodologies are equally effective, as long as a stable reference is identified (classic approach) and the allelic imbalance is caused by the loss or gain/amplification of only one of both alleles (SNP-based approach). Though, heterogeneous copy number alterations can be measured with more precision and sensitivity using the SNP-based approach. In DNA from formalin-fixed and paraffin-embedded samples, the latter approach can also overcome technical artefacts causing inconsistencies between DNA amplicons.

In conclusion, the limits of detecting copy number alterations in heterogeneous DNA can be improved using the SNP-based approach. Consequently, an increasing number of clinical samples may be successfully analysed, providing novel potential for the identification, prognostication and management of cancer.

## Introduction

Cancer develops through the accumulation of somatic genetic alterations over time [1, 2]. For that reason, an adequate and sensitive detection of these alterations is crucial in all phases of cancer care: when the initial diagnosis is made, when a (targeted) therapy may be selected, or during follow-up of tumour progression. These analyses are typically carried out in DNA samples obtained via a tumour or liquid biopsy, and predominantly focus on somatic mutations, small nucleotide changes that may activate oncogenes or inactivate tumour suppressor genes. These mutations typically represent a cancer-cell specific biomarker: the alteration is exclusively present in the DNA of malignant cells, and not in other cells. Regardless of the exact abundance, the qualitative identification of any mutant allele then inevitably indicates that DNA has been analysed that originated from malignant cells bearing this mutation.

Copy number alterations form another relevant group of genetic alterations. Ranging in size from entire chromosomes to only a few kilobases, these alterations are present in up to 90% of cancers [3]. Similar to mutations, the presence of copy number alterations may support the identification of a tumour or its progression, correlates to patient prognosis and is associated with responsiveness to various therapies [3–6]. Though, copy number alterations involve gains or losses of a chromosomal target that is generally present in each cell’s genome. This makes that – in contrast to a mutation – not solely the qualitative identification of this chromosomal target, but an exact quantification is needed to determine any variation in comparison to the copy number neutral state.

A variety of molecular methodologies is available to analyse genetic material and identify copy number alterations [7–11]. In bulk DNA, these alterations are traditionally studied by measuring the relative abundance of a target of interest, normalised against a stable genomic reference (‘classic approach’, **Figure 1A**). This approach forms the basis of various molecular techniques, including genotyping based on arrays or next-generation sequencing, and digital PCR. The latter one involves the compartmentalisation of a PCR reaction into a large number of small partitions (i.e. droplets, crystals or chambers on an array), allowing for an absolute quantification of DNA molecules [8, 12, 13]. By amplifying two amplicons (i.e. the DNA target of interest and a stable reference) and comparing the measured abundances of both, a so-called copy number value can be calculated. This value is defined as the average number of copies of a genomic target per diploid cell. Without an alteration, the copy number value is equal to 2 for autosomal (diploid) DNA targets. When an alteration is present, the value depends on the exact decrease or increase of target copies per cell, and the heterogeneity of the DNA sample (i.e. the fraction of cells in the sample containing the alteration). For example, when 100% of the cells have lost one copy of a diploid target:

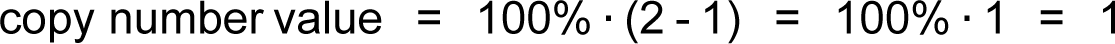

**Figure 1.**
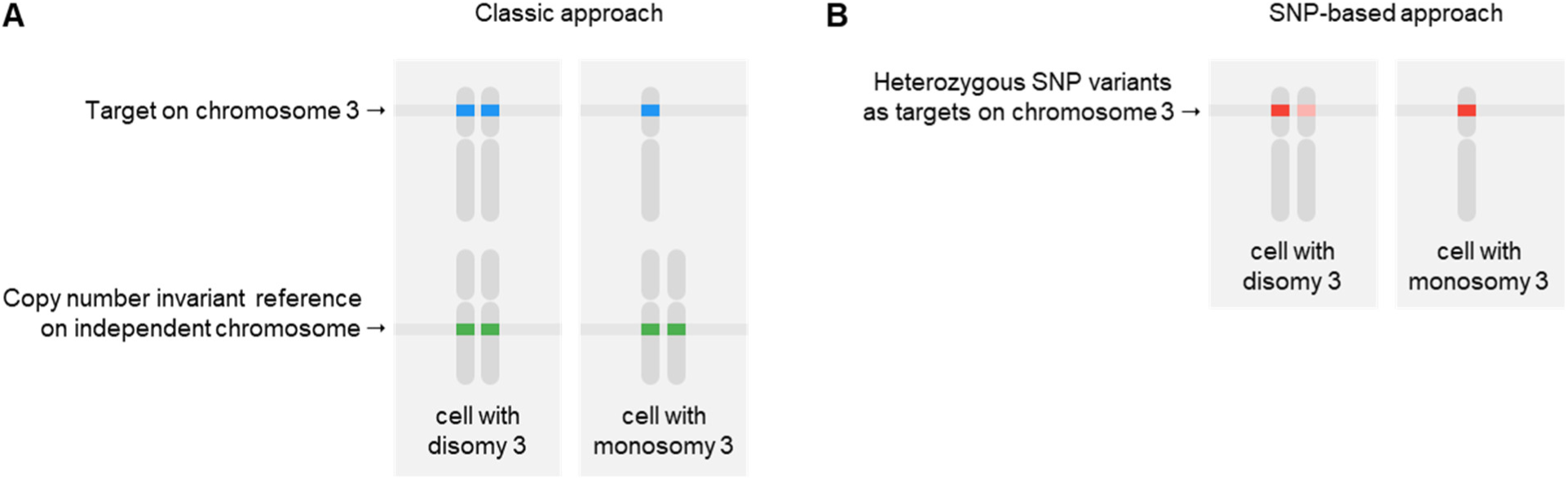
Strategies to analyse copy number alterations, exemplified for measuring a loss of chromosome 3. (**A**) The classic approach involves the comparison of absolute abundances of a target of interest to a copy number invariant, independent genomic reference. (**B**) The alternative SNP-based approach uses the alteration-derived allelic imbalance to detect and quantify the copy number alteration.

When 10% of the cells have gained one copy of such diploid target and the remaining 90% has no alteration:

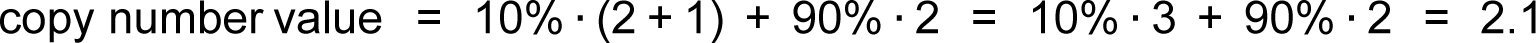

Crucial to the determination of a correct copy number value following this classic approach is the identification of a stable genomic reference gene. This selection is usually guided by information about copy number stability in the malignancy of interest, but an additional analysis is required to exclude sample-specific alterations affecting candidate references [14].

Alternatively, a complementary digital PCR-based approach may be applied to measure copy number alterations without the need of a stable reference gene. This methodology, as we introduced previously [15–17], is based on evaluating the allelic (im)balance between the two variants of a heterozygous single-nucleotide polymorphism (SNP) on the chromosome of interest (‘SNP-based approach’, **Figure 1B**). Whereas both SNP variants are equally abundant under copy number neutral conditions, this balance is usually altered when there is a loss or gain: one of both alleles is deleted or amplified, while the abundance of the other allele remains unaltered. The relative difference in abundance between both alleles can then be used to quantify the copy number alteration.

In this study, we set out to systematically compare the classic and SNP-based experimental setups to measure copy number alterations using digital PCR. Both approaches are modelled *in silico* using a novel R programming library, which is also available as an interactive online application. Next, both methodologies are validated *in vitro* in control samples and in the context of analysing chromosome 3p and 8q copy number alterations in uveal melanoma. Collectively, this study reveals the pitfalls and advantages of both approaches in terms of accuracy, practicability, precision and sensitivity.

## Materials and methods

### Collection and DNA isolation of control samples and uveal melanomas

Two types of copy number neutral control samples from healthy donors were analysed in this study: two samples of freshly isolated peripheral blood mononuclear cells (PBMCs) and two formalin-fixed and paraffin-embedded (FFPE) palatine tonsils [18]. Five snap-frozen primary uveal melanoma specimens were collected from the biobank of the Department of Ophthalmology, Leiden University Medical Center (LUMC). These samples were obtained from patients treated by an enucleation between 2009 and 2010 in the LUMC. This study was approved by the LUMC Biobank Committee and Medisch Ethische Toetsingscommissie under numbers B14.003/SH/sh and B20.026/KB/kb.

DNA from the PBMCs (5 million cells) and uveal melanomas (25x 20 uM sections) was isolated using the QIAmp DNA Mini Kit (Qiagen, Hilden, Germany). The RecoverAll™ Total Nucleic Acid Isolation Kit (Life Technologies, Carlsbad, USA) was used to isolate DNA from the FFPE tonsil specimens (4x 20 uM sections). For all isolations, the respective manufacturer’s instructions were followed.

To evaluate the classic and SNP-based approaches in detecting a heterogeneous copy number alteration, control samples PBMC1 (homozygous for rs1062633) and PBMC2 (heterozygous for rs1062633) were mixed in concentration ratios of respectively 1/40 (classic approach) and 1/20 (SNP-based approach). Consequently, the absolute and relative abundances of both rs1062633 variants were artificially equalled to those of the target and reference genes (following the classic approach), and of var_1_ and var_2_ (following the SNP-based approach) in a DNA sample with a copy number value of 2.1.

### Digital PCR experiments and calculations

We used the QX200 Droplet Digital PCR System (Bio-Rad Laboratories, Hercules, USA) following the general guidelines described and discussed earlier [14, 17, 19, 20]. In summary, 20 ng DNA was analysed in a 22 uL experiment, using 11 uL ddPCR Supermix for Probes (No dUTP, Bio-Rad) and primers and probes in final concentrations of 900 and 250 nM for duplex experiments. These concentrations were optimised for multiplex experiments [21–23]. The context sequence, PCR annealing temperature and supplier information for all assays used is provided in **Supplementary Table 1**.

PCR mixtures were partitioned into 20,000 droplets using the AutoDG System (Bio-Rad). Subsequent PCR was performed in a T100 Thermal Cycler (Bio-Rad) using the following cycle parameters: 10 min at 95 °C; 30 s at 94 °C and 1 min at 55 °C or 60 °C (depending on the assay, see **Supplementary Table 1**) for 40 cycles; 10 min at 98 °C; 30 min at 4 °C; cooling at 12 °C for up to 48 h, until droplet reading. Ramp rate was set to 2 °C/s for all steps. Reading of the droplets was performed using a QX200 Droplet Reader (Bio-Rad).

Raw digital PCR results were acquired using QuantaSoft (version 1.7.4, Bio-Rad) and imported and analysed in the online digital PCR management and analysis application Roodcom WebAnalysis (version 1.9.4, available via https://webanalysis.roodcom.nl).

For the classic approach, copy number values were calculated based on the measured concentrations (listed between square brackets) of the chosen DNA target and one stable diploid reference gene:

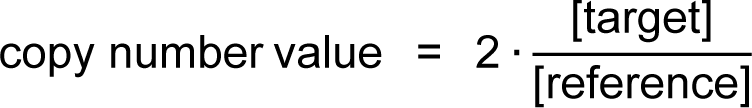

The targets included genes on chromosome 3p (*PPARG*) and 8q (*PTK2*). Genes on chromosome 5p (*TERT*), 7p (*VOPP1*) or 14q (*TTC5*) were evaluated as stable references.

SNP-based copy number values were determined based on the concentrations of haploid genomic loci var_1_ and var_2_, which were the two variants of a heterozygous SNP:

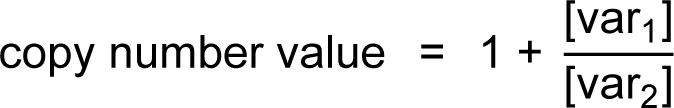

In this formula, the SNP variant listed in the denominator (var_2_) represents the one that remains unaltered (i.e. the haploid copy number stable variant). When measuring a (potential) chromosomal loss (i.e. 3p), the variant with the highest concentration was chosen as stable variant. Vice versa, the variant with the lowest concentration was assumed to be stable when analysing chromosomal gains and amplifications (i.e. 8q). To determine SNP-based copy number values, all samples included in this study were selected for heterozygosity of common SNPs rs1062633 (allele frequencies in the European population according to dbSNP [24], available via https://www.ncbi.nlm.nih.gov/snp: T = 0.50, C = 0.50) or rs2236947 (C = 0.53, A = 0.47) on chromosome 3p, and of SNPs rs7018178 (T = 0.49, C = 0.51) or rs7843014 (A = 0.62, C = 0.38) on chromosome 8q. These SNPs, located in our regions of interest, were chosen because of their high variant allele frequencies, resulting in a high a priori probability of finding heterozygous samples.

For all digital PCR experiments, 99%-confidence intervals were calculated following Fieller’s theorem as described by Dube et al. [8]. *P* values < 0.01 were considered statistically significant.

### Bioinformatic analyses

The workflow of the *in silico* digital PCR experiments is described in **Supplementary Data 1 and 2**. Processed allele-specific copy number data (analysed using ABSOLUTE) from the 80 primary uveal melanomas included in The Cancer Genome Atlas (TCGA) cohort were available from the Pan-Cancer Atlas initiative [3, 25, 26]. For the analyses, R (version 4.0.3), RStudio (version 1.4.1103) and R packages rmarkdown (version 2.7) and digitalPCRsimulations (version 1.1.1, available via https://github.com/rjnell/digitalPCRsimulations) were used.

## Results

### Development and validation of an R library to perform *in silico* simulations of digital PCR experiments

To evaluate the stochastic variability and precision of digital PCR experiments *in silico*, we developed the R programming library ‘digitalPCRsimulations’. This library is based on earlier work [8, 15, 27] and follows the key principles of *in vitro* experiments. The workflow is described in detail in **Supplementary Data 1** and summarised in **Figure 2A**. In short, digital PCR experiments are simulated by creating large virtual universes of droplets, which are filled randomly with target molecules based on the number of target copies in a given amount of input DNA. Next, a random sample of droplets is taken and the number of positive (i.e. target-containing) droplets is measured and interpreted mathematically in a similar way as the result of an *in vitro* digital PCR experiment. With only a fraction of the universe of droplets being measured, subsampling of the DNA sample is also mimicked *in silico*.

**Figure 2.**
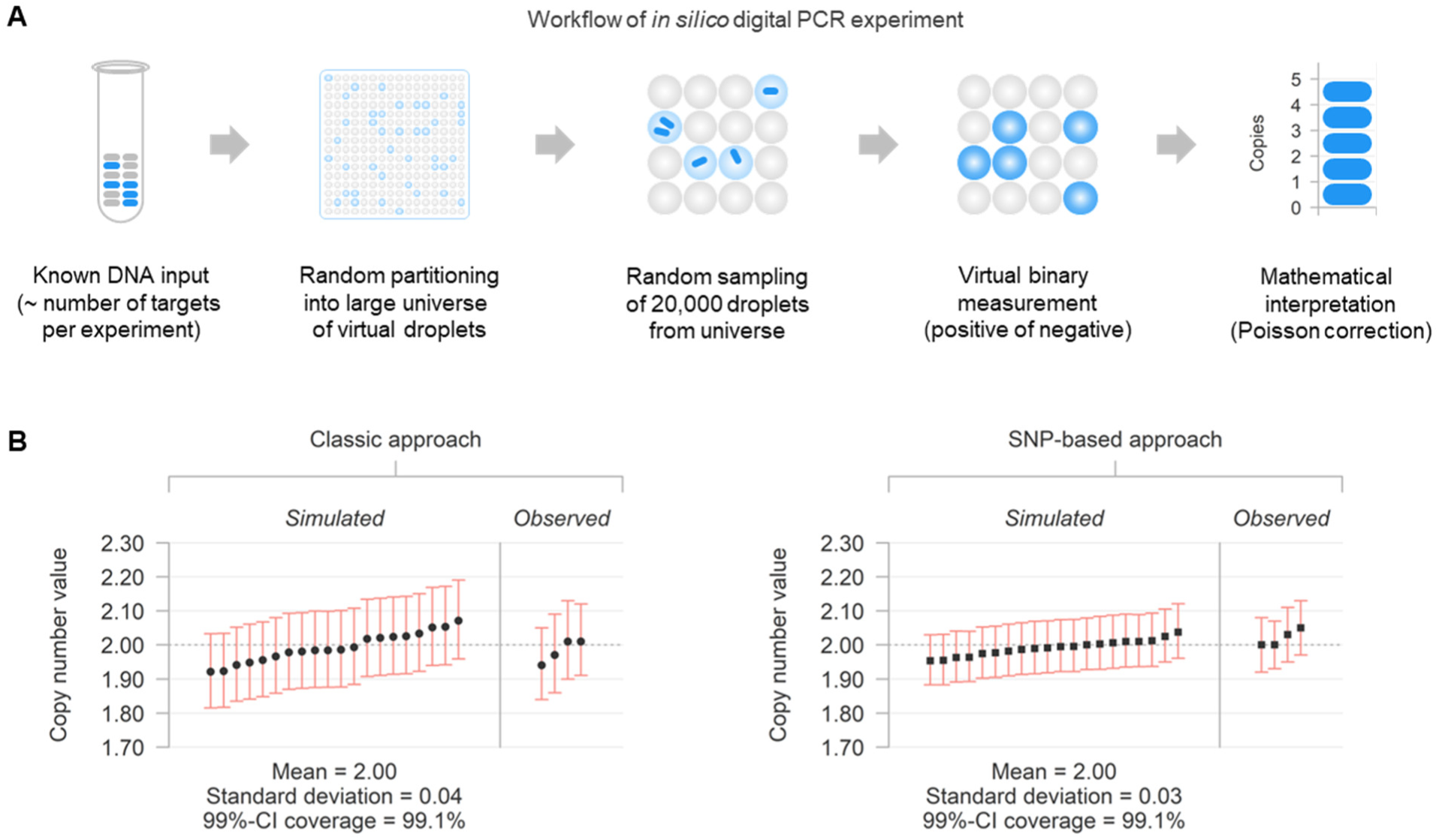
*In silico* and *in vitro* digital PCR to determine copy number values according to the classic and SNP-based approach. (**A**) Workflow of *in silico* digital PCR experiments, consisting of virtual partioning and sampling of input targets with subsequent measurement and mathematical analysis. (**B**) Comparison of copy number values determined by the classic and SNP-based approaches using *in silico* and *in vitro* digital PCR experiments in 20 ng copy number invariant DNA (true copy number value = 2, as indicated by the grey dotted line). The individual results of the first 20 *in silico* and all 4 *in vitro* experiments per approach are sorted and plotted with accompanying 99%-confidence interval (CI). The statistics (mean, standard deviation and fraction of the CIs containing the true copy number value [coverage]) from all simulations (n = 10,000) are presented below the plots.

To validate the *in silico* experiments, we simulated the analysis of 20 ng copy number neutral genomic DNA and compared the outcomes with those from similar *in vitro* experiments measuring healthy PBMC DNA (**Figure 2B**). Following the classic approach, copy number values were determined based on the concentrations of independent diploid genomic loci ‘target’ and ‘reference’. SNP-based copy number values were determined based on the concentrations of haploid genomic loci ‘var_1_’ and ‘var_2_’, which are the two alleles of a heterozygous SNP. To indicate the uncertainty of individual measurements, a 99%-confidence interval was calculated for each experiment following Fieller’s theorem [8]. Overall, highly similar results were obtained when comparing the *in silico* and *in vitro* results (**Figure 2B**), which validated that real digital PCR experiments can be adequately mirrored *in silico*.

The mean of the determined copy number values was close to the true value of 2 in all experiments, which also confirmed the validity of the classic and SNP-based model. Moreover, the 99%-confidence intervals for all *in silico* experiments contained the true copy number value in a fraction of the experiments very close to the intended 99%, indicating that that – under these neutral conditions – the construction of the confidence intervals was mathematically adequate. Importantly, however, the standard deviations of the copy number values and absolute width of the confidence intervals were smaller when using the SNP-based approach. This was evident from both the *in silico* and *in vitro* experiments (**Figure 2B**).

### *In silico* and *in vitro* evaluation of detecting copy number alterations

As a further comparison of the two digital PCR experimental setups, we performed additional *in silico* and *in vitro* experiments for a variety of heterogeneous DNA mixtures. First of all, we extended the simulations to copy number values between 1.0 and 9.0 (with intervals of 0.1) in single experiments with 20 ng genomic input DNA. Again, both approaches resulted in very comparable mean copy number values close to the expected values, but with varying standard deviations (**Figure 3A and B**). To determine the precision of the classic and SNP-based methodologies, the coefficient of variant was calculated for all simulated conditions (**Figure 3C**). This analysis showed that – for copy number values < 4.6 – the SNP-based approach was more precise: as evidenced by the lower coefficient of variation, the determined values were closer to the true values than when using the classic approach.

**Figure 3.**
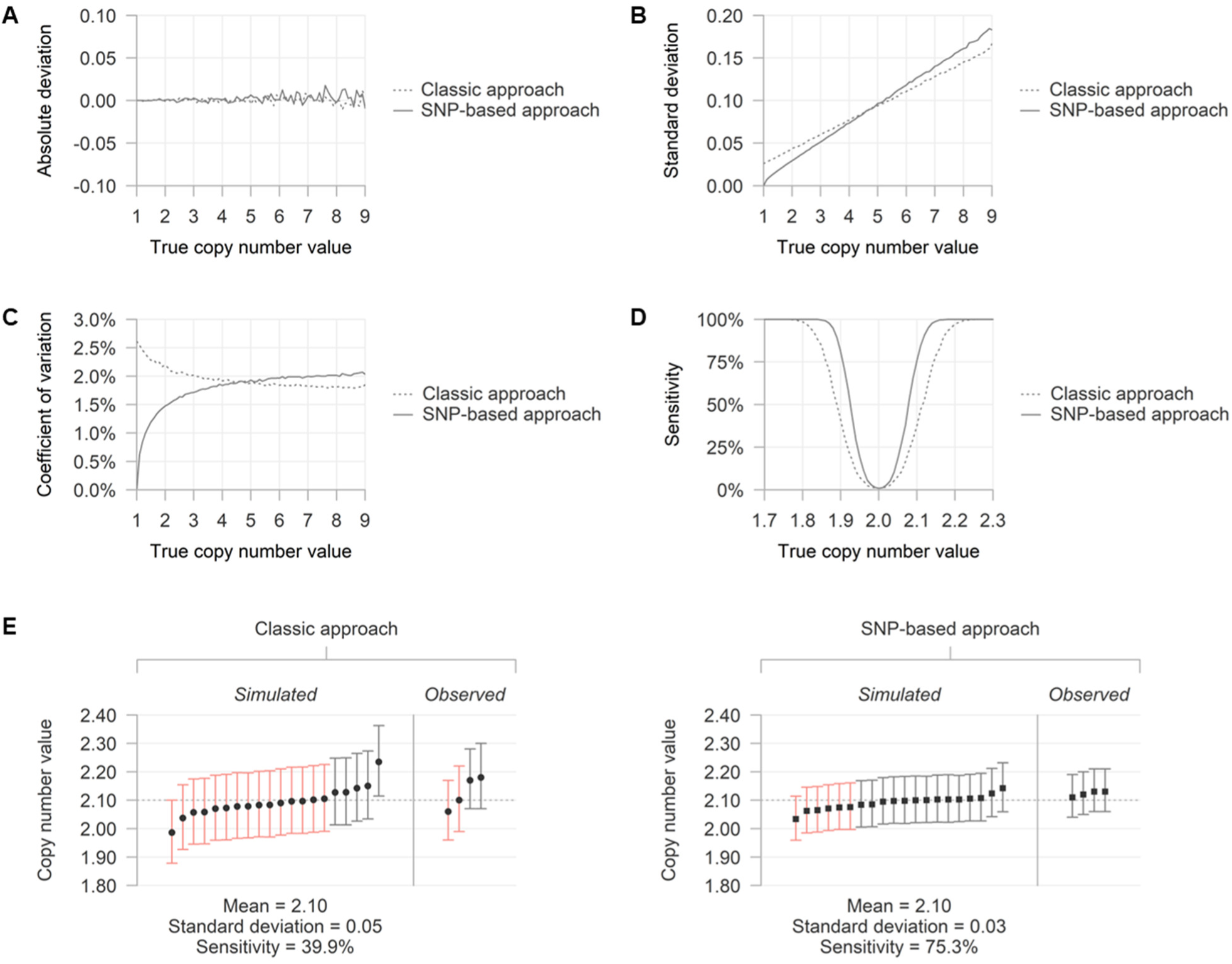
Performance of classic and SNP-based approaches in detecting and quantifying heterogeneous copy number alterations. All conditions are evaluated *in silico* using 10,000 simulations with 20 ng DNA and 17,500 droplets. (**A**) Absolute deviation, defined as the difference between the mean of measured copy number values minus the true copy number value, per simulated condition. (**B**) Standard deviation of measured copy number values per simulated condition. (**C**) Coefficient of variation of measured copy number values per simulated condition. (**D**) Sensitivity of detecting a copy number alteration, defined as the percentage experiments with an outcome significantly different from 2, per simulated condition. (**E**) Comparison of copy number values determined by the classic and SNP-based approaches using *in silico* and *in vitro* digital PCR experiments in 20 ng copy number altered DNA (true copy number value = 2.1, as indicated by the grey dotted line). The results of the first 20 *in silico* and all 4 *in vitro* experiments per approach are sorted and plotted with accompanying 99% confidence interval (CI). Significance of the copy number gains is indicated by the colour of the individual CI (red: not significantly different from 2 [*p* > 0.01], grey: significantly different from 2 [*p* < 0.01]). The statistics (mean, standard deviation and fraction of the CIs containing the true copy number value [coverage]) from all simulations (n = 10,000) are presented below the plots.

Secondly, we evaluated whether the 99%-confidence intervals were adequately determined in the simulated experiments. Indeed, in all conditions, ∼99% of the intervals contained the true copy number value, confirming their correctness. This *in silico* analysis has potential translational relevance: as in a single experiment a copy number alteration can only be considered ‘present’ when the result is significantly different from 2 (i.e. the confidence interval does not contain 2), simulations may be used to determine the statistical power to call a copy number alteration. In other words: given a certain condition, in how many of the experiments may a significant result be expected?

To answer this question, we simulated copy number values close to 2 and evaluated the proportion experiments with an outcome significantly different from 2 (i.e. the sensitivity of detecting an alteration, **Figure 3D**). In line with our earlier results, higher levels of sensitivity were observed when using the SNP-based approach. For example, when analysing copy number value of 2.1 (e.g. derived from 10% trisomic and 90% disomic cells) in 20 ng of DNA, this alteration was only detected in 39.9% of the simulated classic experiments, compared to 75.3% of the SNP-based experiments (**Figure 3D**). To confirm these observations *in vitro*, control samples were mixed to generate a heterogenous DNA sample with a measurable copy number value of 2.1. Indeed, more precise and smaller confidence intervals were obtained with the SNP-based approach, leading to more experiments that had a significantly detectable alteration (**Figure 3D**).

The precision and sensitivity of both methodologies were also evaluated with other amounts of input DNA and total numbers of droplets analysed (**Supplementary** Figure 1). Although the absolute differences between the two approaches varied, the advantages of the SNP-based approach were consistently present throughout the simulated conditions. In addition to our R library (see **Supplementary Data 1** for all scripts to reproduce the simulations of this manuscript), an interactive online application is available via https://www.roodcom.nl/digitalPCRsimulations.php to perform simulations of both approaches for adjustable individual input conditions.

### Evaluation of chromosome 3p and 8q copy number alterations in uveal melanoma

To further test the performance of both the classic and SNP-based approaches *in vitro*, we studied two recurrent copy number alterations in primary uveal melanoma specimens. In this malignancy, the (arm-level) loss of chromosome 3p and gain or amplification of chromosome 8q are the most prevalent and prognostically-relevant chromosomal alterations [19, 20, 28]. Within the 80 primary uveal melanomas studied by TCGA initiative using bulk genotyping [26], 48% and 70% of the tumours carried a chromosome 3p or 8q alteration, respectively (**Figure 4A**). This contrasts with the much lower frequencies that loci 5p (5%), 7p (11%) and 14q (10%) were involved in a copy number alteration, warranting the usage of these chromosomal regions as candidate references in digital PCR experiments following the classic approach, as previously published [17, 19, 20, 29].

**Figure 4.**
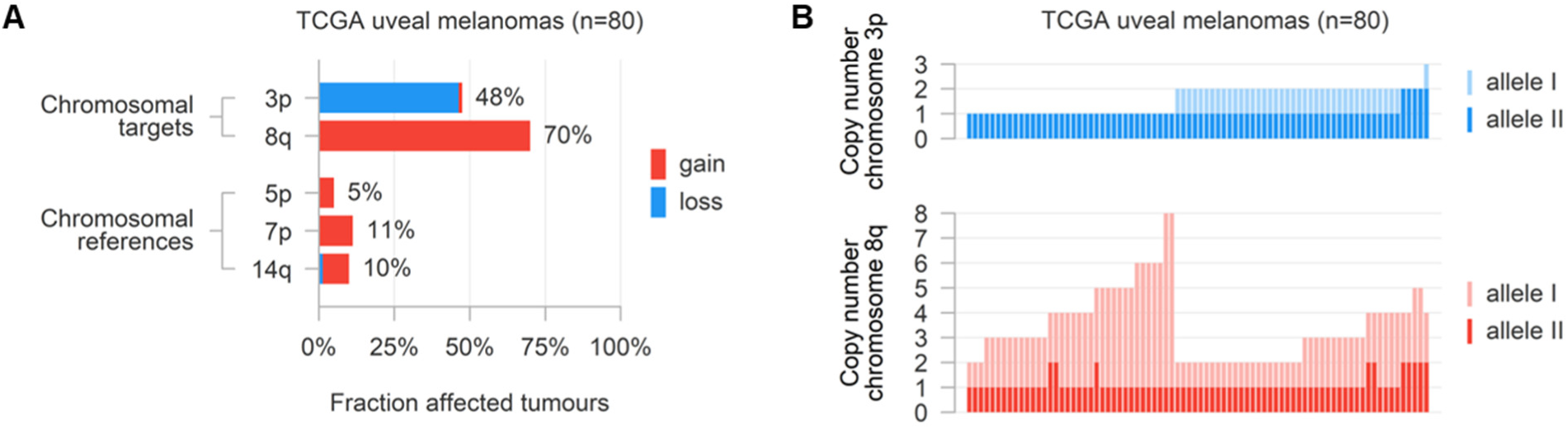
Copy number alterations in 80 primary uveal melanomas studied via bulk genotyping by TCGA. (**A**) Frequencies of chromosomal losses and gains involving target loci 3p or 8q and candidate reference loci 5p, 7p and 14q. (**B**) Modal allele-specific copy numbers of chromosomes 3p and 8q.

To gain insight into the allele-specificity of chromosome 3p and 8q alterations in uveal melanoma, we also evaluated the modal allelic copy number of all TCGA primary tumours (**Figure 4B**). Overall, most affected cases showed a copy number alteration consisting of the loss or gain/amplification of only one of both alleles. Only 5/80 (6%) tumours showed an unbalanced disomy 3p (i.e. isodisomy, n=4) or unexpected gain of chromosome 3p (n=1), and 10/80 (13%) tumours demonstrated a biallelic amplification of chromosome 8q. Extrapolating these results to our digital PCR-based application, respectively 94% and 88% of the chromosome 3p and 8q copy number values would be analysed correctly using the SNP-based approach.

To evaluate the classic and SNP-based approaches *in vitro*, we selected five primary uveal melanomas (UM-1 to UM-5) from our own cohort to be analysed using digital PCR. A representative example of the analysis is shown in **Figure 5** and the results of all analyses are summarised in **Table 1**. To quantify copy number alterations following the classic approach, the abundance of the 3p and 8q target genes was measured against one stable genomic reference gene. Reference stability was ensured by comparing the three candidate reference genes on chromosome 5p, 7p and 14q in a separate (classic) digital PCR experiment. For this evaluation, we developed a 1×2 multiplex setup, in which the three candidates were measured concurrently (**Figure 5A**). In most melanomas, the measured abundances of the references were non-significantly different from each other and either gene could be arbitrarily selected as a representative, stable reference (**Figure 6A**). In UM-4, however, a decreased abundance of chromosome 14q compared to the other two candidate references suggested a copy number loss affecting this chromosomal region. Therefore, 14q was not considered stable in this tumour. Since in all samples either chromosome 5p or 14q was selected to be stable, we combined these two references with our targets 3p and 8q in a 2×2 multiplex setup to calculate copy number values following the classic approach (**Figure 5B**). With this generic multiplex, all uveal melanomas could be analysed using the same experimental setup, while in individual samples only one reference (the one considered stable) was utilised in the downstream calculations.

**Table 1.** Summary of copy number results with accompanying 99%-confidence interval (CI) in all PBMCs, uveal melanomas and tonsils.

| Sample |  | Reference stability |  |  | Copy number value chromosome 3p (99%-CI) |  | Copy number value chromosome 8q (99%-CI) |  |
| --- | --- | --- | --- | --- | --- | --- | --- | --- |
|  |  | 5p | 7p | 14q | Classic approach | SNP-based approach | Classic approach | SNP-based approach |
| PBMCs | PBMC-1 | = | = | = | 1.97 (1.86-2.09) | 2.00 (1.92-2.08) | 2.01 (1.90-2.13) | 2.03 (1.95-2.11) |
|  | PBMC-2 | = | = | = | 1.95 (1.84-2.06) | 2.00 (1.96-2.07) | 2.02 (1.91-2.13) | 2.02 (1.96-2.10) |
| Uveal melanomas | UM-1 | = | = | = | 1.16 (1.08-1.24) | 1.17 (1.15-1.20) | 3.48 (3.30-3.67) | 3.34 (3.19-3.51) |
|  | UM-2 | = | = | = | 2.11 (1.99-2.23) | 2.00 (1.93-2.07) | 2.00 (1.89-2.11) | 2.01 (1.93-2.10) |
|  | UM-3 | = | = | = | 1.22 (1.13-1.31) | 1.17 (1.15-1.20) | 2.10 (1.97-2.23) | 2.12 (2.04-2.20) |
|  | UM-4 | = | = | ↓ | 1.20 (1.11-1.29) | 1.18 (1.16-1.22) | 6.34 (6.00-6.71) | 6.16 (5.86-6.48) |
|  | UM-5 | = | = | = | 1.39 (1.30-1.49) | 1.45 (1.41-1.50) | 3.08 (2.90-3.27) | 3.09 (2.93-3.26) |
| Tonsils | T-1 | = | ↑ | = | 1.69 (1.58-1.80) | 1.95 (1.87-2.03) | 2.14 (2.01-2.28) | 2.05 (1.97-2.15) |
|  | T-2 | ↓ | = | = | 1.97 (1.88-2.08) | 1.84 (1.66-2.04) | 2.08 (1.98-2.18) | 2.01 (1.93-2.10) |

no significant alteration loss ( $p < 0.01$ ) gain ( $p < 0.01$ )

**Figure 5.**
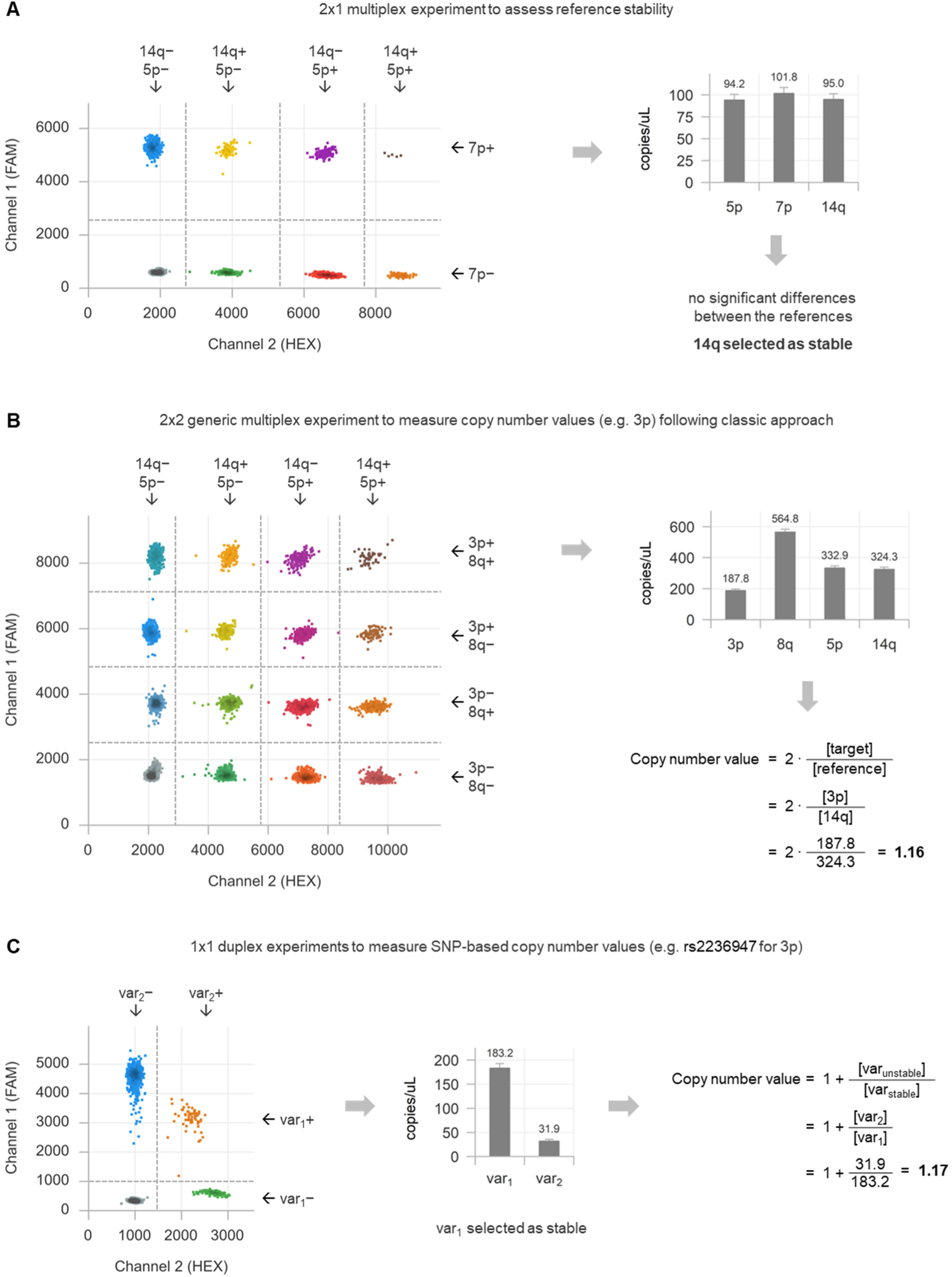
Representative example (of UM-1) of complete copy number value analyses using the classic and SNP-based approach. (**A**) Two-dimensional plot of 1×2 multiplex digital PCR experiment measuring the abundances of candidate references on chromosome 7p (single assay on channel 1), 5p (assay with lowest amplitude on channel 2) and 14q (assay with highest amplitude on channel 2), with associated outcomes and interpretation of reference stability. (**B**) Two-dimensional plot of 2×2 multiplex digital PCR experiment measuring the abundances of targets on chromosome 8q (assay with lowest amplitude on channel 1) and 3p (assay with highest amplitude on channel 1), and candidate references on chromosome 5p (lowest amplitude on channel 2) and 14q (assay with highest amplitude on channel 2), with associated outcomes and interpretation of the copy number value according to the classic approach of chromosome 3p. (**C**) Two-dimensional plot of 1×1 duplex digital PCR experiment measuring the abundances of the two variants of common SNP rs2236947 on chromosome 3p, with associated outcomes and interpretation of the SNP-based copy number value of chromosome 3p.

**Figure 6.**
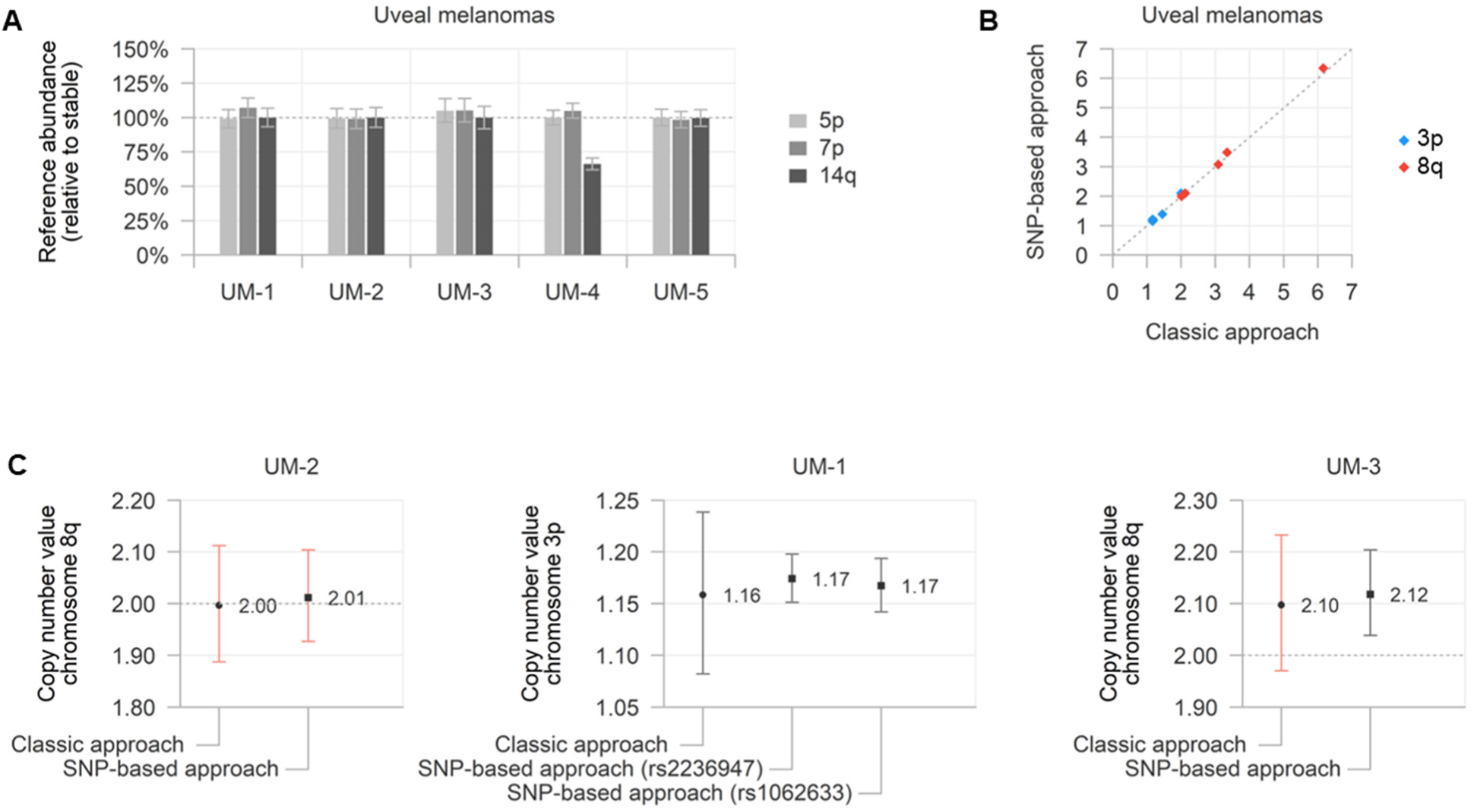
Results of analyses in five primary uveal melanoma specimens (UM-1 to UM-5). (**A**) Measured abundances of the three candidate references on chromosome 5p, 7p and 14q with accompanying 99%-confidence interval (CI), relative to the selected stable reference. (**B**) Comparison between copy number values determined by the classic and SNP-based approaches for chromosome 3p and 8q in all uveal melanomas, see also **Table 1**. (**C**) Examples of copy number values with accompanying 99%-CI determined by the two approaches in three tumours. Significance of the copy number alterations is indicated by the colour of the individual CI (red: not significantly different from 2 [*p* > 0.01], grey: significantly different from 2 [*p* < 0.01]).

Next, the two variants of heterozygous SNPs on chromosome 3p and 8q were quantified using digital PCR to measure the allelic imbalances (**Figure 5C**). For this goal, all five uveal melanomas were selected for heterozygosity of common SNPs on chromosome 3p (rs1602633 or rs2236947) and chromosome 8q (rs7018178 or rs7843014).

The copy number values determined by both approaches in the five uveal melanomas are presented in **Figure 6B** **and Table 1**. The high concordance (Spearman’s rho = 0.95, *p* < 0.001) between both measurements cross-validated our experimental set-ups. UM-1 presented with heterozygosity of both chromosome 3p SNPs: the high similarity of the two distinct SNP measurements confirmed the technical reproducibility between both assays (**Figure 6C**). Moreover, in most samples the SNP-based copy number values showed smaller confidence intervals (**Figure 6C**). Most notably, in UM-3 a small 8q copy number increase was only detected by the SNP-based approach: there was a significant allelic imbalance, but the difference between target and reference following the classic approach did not reach statistical significance.

### Evaluation of copy number stability in FFPE-derived healthy tonsils

Whereas our previous DNA samples were isolated from fresh PBMCs or frozen melanoma tissues, a substantial part of all clinical samples originates from FFPE specimens. Though, FFPE-derived DNA samples are generally more challenging to analyse, as they are known to be (differentially) degraded and affected by chemical artefacts [30–32]. To evaluate the performance of the classic and SNP-based approaches in FFPE-derived DNA, we analysed two specimens obtained from palatine tonsils. These tissues are of non-neoplastic origin and can therefore be considered biologically copy number invariant. Consequently, no copy number alterations are expected and any detected alteration must originate from a technical artefact.

Both tonsils were analysed following the same experimental procedures as the uveal melanoma samples (illustrated in **Figure 5**), and the results are summarised in **Table 1**. Notably, in all experiments the thresholds were more difficult to set due to the presence of ‘rain’ (**Figure 7A**). Comparing the candidate reference genes, unexpected copy number variation was observed in both tonsils T-1 (gain of chromosome 7p) and T-2 (loss of chromosome 5p, **Figure 7B**). Moreover, chromosome 3p and 8q were not entirely copy number stable when determined by the classic approach (**Figure 7C**). The observed alterations were not consistently present between both samples. In contrast, heterozygous SNPs on chromosome 3p and 8q did not show allelic imbalances and all SNP-based copy number values were not significantly different from 2 (**Figure 7D**).

**Figure 7.**
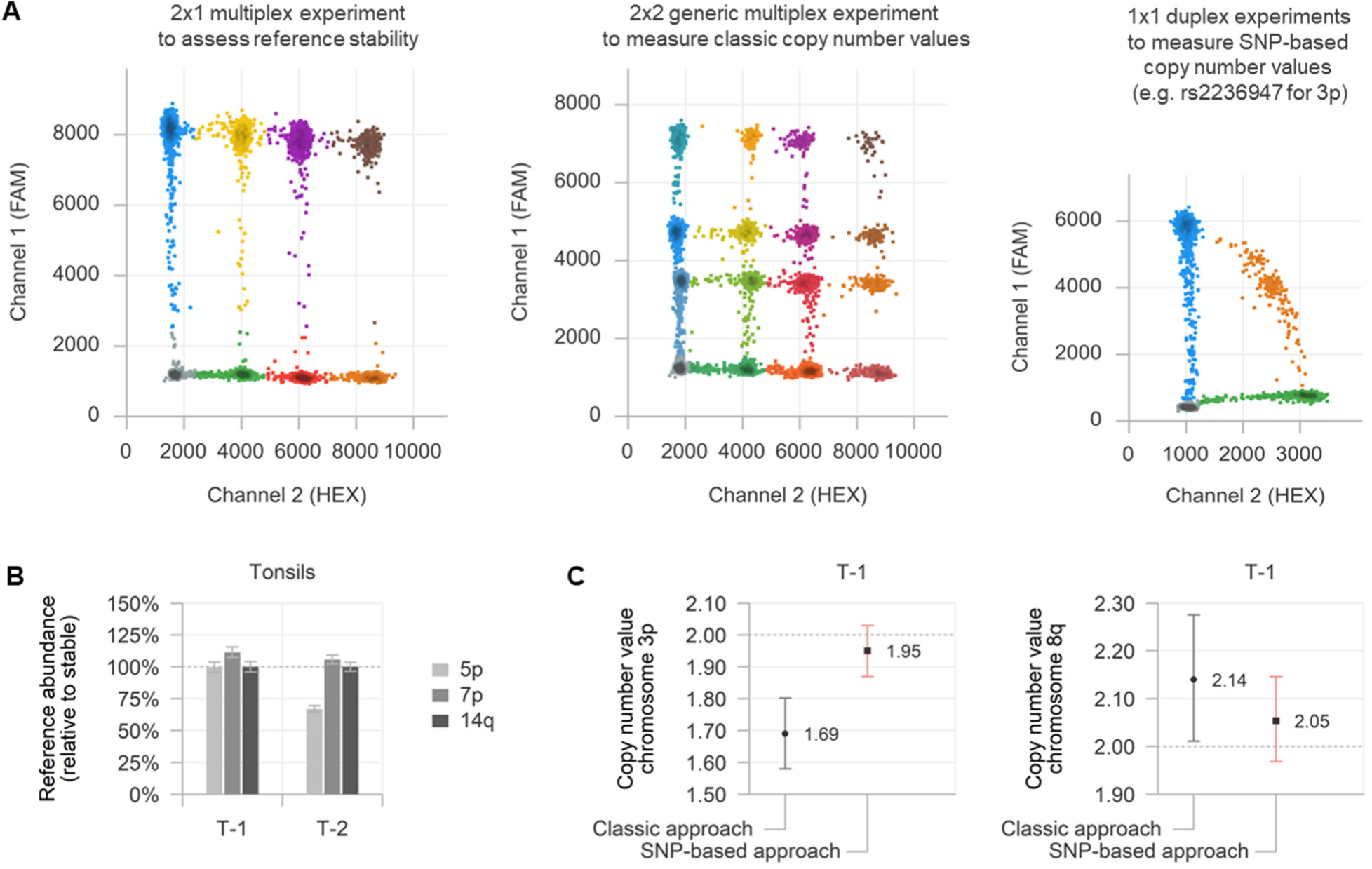
Results of analyses in two FFPE-derived biologically copy number stable tonsil specimens (T-1 and T-2). (**A**) Representative two-dimensional plots (of T-1) of multiplex and duplex experiments, demonstrating clustering difficulties (compared to same experiments in non-FFPE derived samples from Figure 5). (**B**) Measured abundances of the three candidate references on chromosome 5p, 7p and 14q with accompanying 99%-confidence interval (CI), relative to the selected stable reference. (**C**) Chromosome 3p and 8q copy number values with accompanying 99%-CI as determined by the two approaches in tonsil T-1, see also **Table 1**. Significance of the copy number alterations is indicated by the colour of the individual CI (red: not significantly different from 2 [*p* > 0.01], grey: significantly different from 2 [*p* < 0.01]).

## Discussion

During the development and evolution of cancer, a variety of somatic mutations and copy number alterations accumulate which impact the aggressiveness of the disease, but also influence potential treatment response or resistance [1–6, 33, 34]. An accurate and sensitive detection of the genetic alterations present in a tumour is – given these reasons – of vital importance in cancer care. However, tumour material is usually only scarcely available and almost never entirely pure and homogeneous.

In contrast to mutations, in which the qualitative detection of any mutant allele evidences their presence, copy number alterations can only be detected as quantitative variation in comparison to the copy number neutral state. Consequently, these alterations are technically more difficult to measure and this particularly applies to samples where the alteration is present in only a fraction of the DNA: ‘contaminating’ DNA from invariant cells complicates the detection of the particular alteration. As a result, copy number alterations are less frequently analysed in heterogeneous clinical samples of lower purity or quantity, such as small solid or liquid tumour biopsies [35].

When interested in a targeted, deep quantitative analysis of DNA samples, digital PCR forms one of the most accessible and most successful technologies in the field [36]. In the current study, we explored and validated two complementary digital PCR-based approaches to detect and quantify copy number alterations. The accuracy, practicability, precision and sensitivity of both methodologies were evaluated extensively *in silico* using a newly introduced R library, and *in vitro* by analysing copy number (in)variant control samples and malignant uveal melanoma specimens.

Using digital PCR, copy number alterations are traditionally measured by comparing the absolute abundances of a chromosomal target of interest with a genomically stable reference located on another chromosome. Although this ‘classic’ approach seems relatively generic, the identification of such copy number invariant reference gene can be extremely challenging, especially in heterogeneous, genomically unstable tumour material. As a pragmatic solution, we measured a variety of candidate references in a separate multiplex digital PCR experiment to ensure the absence of variability of the chosen reference gene. Though, despite selecting potential references based on their renowned copy number stability in the malignancy of interest, unexpected sample-specific (subclonal) alterations may always be present. Identification of the correct reference is therefore only possible after rigorous testing of each individual tumour specimen. This procedure requires additional experiments and is sample-expensive, providing a barrier to the proper analysis of potentially unstable DNA specimens of lower quantity.

As an orthogonal alternative, copy number alterations may be measured by quantifying the allelic imbalances that underlie these alterations. We determined these imbalances by analysing the abundances of both variants of a heterozygous (common) SNP present in the locus of interest, using a dedicated digital PCR assay. Advantageously, this approach does not require an independent genomic reference: as usually only one of both alleles is gained or lost, the other, unaltered allele functions as (haploid) copy number stable reference. The identification of a sample-specific heterozygous SNP may be guided by known variant allele frequencies in the population of interest and then directly measured in tumour DNA, or may be carried out by testing germline DNA (e.g. saliva, blood) from individual patients, which is usually less precious than tumour material. As an additional advantage, we found that the SNP-based approach can be more precise than the classic approach, while requiring the same amount of input DNA (**Figure 2B, 3 and 6C**). Though this increased precision depends on various parameters (such as the true copy number value, the exact amount of input DNA and the total number of droplets analysed, **Figure 3** **and Supplementary** Figure 1), it can result in markedly and clinically-relevant higher levels of sensitivity, especially when measuring chromosomal losses or gains in heterogeneous DNA samples (**Figure 3 and 6C**).

Strikingly, we also observed inconsistent technical biases leading to incorrect copy number values when analysing FFPE-derived DNA samples following the classic approach, which was not evident when using the SNP-based approach (**Figure 7**). We hypothesise that FFPE-derived DNA contains various levels of degradation and fragmentation across different genomic loci, that may lead to amplicon-dependent differences in PCR availability (and consequent measurability) which may vary between individual samples. Similar observations have been made in other analyses of FFPE-derived DNA samples [16, 30–32]. These differences may be incorrectly interpreted as a copy number alteration when comparing the measured abundances of DNA targets from different amplicons (i.e. the classic approach). In contrast, this bias may be overcome by using the SNP-based approach: the two variants of a heterozygous SNP are analysed from only a single amplicon and unexpected imbalances were not observed. Given these observations, special care should be taken when analysing FFPE-derived DNA samples for copy number alterations using digital PCR.

The classic and SNP-based methodology were evaluated *in silico* by our R library *digitalPCRsimulations*, which is also available as an interactive online application (https://www.roodcom.nl/digitalPCRsimulations). We showed that using these simulations, the outcomes of digital PCR experiments and the sensitivity (‘power’) of copy number alterations based on a given input condition could be adequately predicted. A potential application of this workflow may be found in the analysis of heterogenous material with only a minor proportion of the DNA containing a particular copy number alteration, such as (liquid) biopsies: based on the input DNA concentration and expected sample purity, the sensitivity of detecting an alteration may be evaluated before doing the actual experiment. Whenever the predicted sensitivity is too low, a researcher might decide not to perform the experiment, by which the number of false-negative results can be reduced and scarce DNA material may be saved. On the other hand, the *in silico* experiments could also be used to determine how this sensitivity may be increased, for example by using the SNP-based approach instead of the classic approach, or by repeating the experiment (i.e. increasing the total number of analysed droplets) and merging the obtained results.

Nevertheless, it is important to note the various technical and biological causes that may hamper the exact quantification of copy number alterations. As mentioned before, a stable reference for the classic approach may be hard to be identified in highly unstable or (heterogeneously) polyploid samples. On the other hand, when no heterozygous SNP is available in the target locus of interest, or when an alteration involves both alleles (e.g. a biallelic amplification or isodisomy), the exact copy number value cannot be determined by solely analysing the allelic imbalance. Also, when an alteration involves the loss or gain of only one of both alleles, but the direction of the alteration is unknown, the SNP-based approach may still identify an imbalance, but might be insufficient to determine the exact copy number value. After all, combining both methodologies would be the preferred and most complete solution when DNA quality and quantity allow.

In conclusion, we technically and biologically validated two complementary digital PCR-based approaches for analysing copy number alterations in heterogeneous DNA mixtures. While requiring an allele-specific imbalance, the SNP-based approach can measure chromosomal losses and gains/amplifications with superior precision and sensitivity, is less prone to FFPE-derived biases and does not require an independent genomic reference gene. Consequently, the limits of detecting copy number alterations in heterogeneous DNA can be improved and an increasing number of clinical samples may be successfully analysed, providing novel potential for the identification, prognostication and management of cancer.

## Data Availability

All data produced in the present work are contained in the manuscript.

https://github.com/rjnell/digitalPCRsimulations

## Acknowledgements

Supported by the European Union’s Horizon 2020 Research and Innovation Program under grant agreement number 667787 (UM Cure 2020 project; Rogier J. Nell).

## References

1. Martincorena, I. and P.J. Campbell, Somatic mutation in cancer and normal cells. Science, 2015. 349(6255): p. 1483–9.

2. Jolly, C. and P. Van Loo, Timing somatic events in the evolution of cancer. Genome Biol, 2018. 19(1): p. 95.

3. Taylor, A.M., et al., Genomic and Functional Approaches to Understanding Cancer Aneuploidy. Cancer Cell, 2018. 33(4): p. 676–689 e3.

4. Davoli, T., et al., Tumor aneuploidy correlates with markers of immune evasion and with reduced response to immunotherapy. Science, 2017. 355(6322).

5. Hieronymus, H., et al., Tumor copy number alteration burden is a pan-cancer prognostic factor associated with recurrence and death. Elife, 2018. 7.

6. Ben-David, U. and A. Amon, Context is everything: aneuploidy in cancer. Nat Rev Genet, 2020. 21(1): p. 44–62.

7. Pinkel, D., et al., High resolution analysis of DNA copy number variation using comparative genomic hybridization to microarrays. Nat Genet, 1998. 20(2): p. 207–11.

8. Dube, S., J. Qin, and R. Ramakrishnan, Mathematical analysis of copy number variation in a DNA sample using digital PCR on a nanofluidic device. PLoS One, 2008. 3(8): p. e2876.

9. Yoon, S., et al., Sensitive and accurate detection of copy number variants using read depth of coverage. Genome Res, 2009. 19(9): p. 1586–92.

10. Pinto, D., et al., Comprehensive assessment of array-based platforms and calling algorithms for detection of copy number variants. Nat Biotechnol, 2011. 29(6): p. 512–20.

11. Talevich, E., et al., CNVkit: Genome-Wide Copy Number Detection and Visualization from Targeted DNA Sequencing. PLoS Comput Biol, 2016. 12(4): p. e1004873.

12. Vogelstein, B. and K.W. Kinzler, Digital PCR. Proc Natl Acad Sci U S A, 1999. 96(16): p. 9236–41.

13. Hindson, B.J., et al., High-throughput droplet digital PCR system for absolute quantitation of DNA copy number. Anal Chem, 2011. 83(22): p. 8604–10.

14. Zoutman, W.H., R.J. Nell, and P.A. van der Velden, Usage of Droplet Digital PCR (ddPCR) Assays for T Cell Quantification in Cancer. Methods Mol Biol, 2019. 1884: p. 1–14.

15. Nell, R.J., et al., Quantification of DNA methylation independent of sodium bisulfite conversion using methylation-sensitive restriction enzymes and digital PCR. Hum Mutat, 2020. 41(12): p. 2205–2216.

16. Christodoulou, E., et al., Loss of Wild-Type CDKN2A Is an Early Event in the Development of Melanoma in FAMMM Syndrome. J Invest Dermatol, 2020. 140(11): p. 2298–2301 e3.

17. Nell, R.J., et al., Involvement of mutant and wild-type CYSLTR2 in the development and progression of uveal nevi and melanoma. BMC Cancer, 2021. 21(1): p. 164.

18. Zoutman, W.H., et al., Accurate Quantification of T Cells by Measuring Loss of Germline T-Cell Receptor Loci with Generic Single Duplex Droplet Digital PCR Assays. J Mol Diagn, 2017. 19(2): p. 236–243.

19. Versluis, M., et al., Digital PCR validates 8q dosage as prognostic tool in uveal melanoma. PLoS One, 2015. 10(3): p. e0116371.

20. de Lange, M.J., et al., Heterogeneity revealed by integrated genomic analysis uncovers a molecular switch in malignant uveal melanoma. Oncotarget, 2015. 6(35): p. 37824–35.

21. Zhong, Q., et al., Multiplex digital PCR: breaking the one target per color barrier of quantitative PCR. Lab Chip, 2011. 11(13): p. 2167–74.

22. Whale, A.S., J.F. Huggett, and S. Tzonev, Fundamentals of multiplexing with digital PCR. Biomol Detect Quantif, 2016. 10: p. 15–23.

23. Nell, R.J., et al., Generic Multiplex Digital PCR for Accurate Quantification of T Cells in Copy Number Stable and Unstable DNA Samples. Methods Mol Biol, 2022. 2453: p. 191–208.

24. Sherry, S.T., M. Ward, and K. Sirotkin, dbSNP-database for single nucleotide polymorphisms and other classes of minor genetic variation. Genome Res, 1999. 9(8): p. 677–9.

25. Carter, S.L., et al., Absolute quantification of somatic DNA alterations in human cancer. Nat Biotechnol, 2012. 30(5): p. 413–21.

26. Robertson, A.G., et al., Integrative Analysis Identifies Four Molecular and Clinical Subsets in Uveal Melanoma. Cancer Cell, 2017. 32(2): p. 204–220 e15.

27. Nell, R.J., et al., Accurate Quantification of T Cells in Copy Number Stable and Unstable DNA Samples Using Multiplex Digital PCR. J Mol Diagn, 2022. 24(1): p. 88–100.

28. Horsman, D.E. and V.A. White, Cytogenetic analysis of uveal melanoma. Consistent occurrence of monosomy 3 and trisomy 8q. Cancer, 1993. 71(3): p. 811–9.

29. de Lange, M.J., et al., Digital PCR-Based T-cell Quantification-Assisted Deconvolution of the Microenvironment Reveals that Activated Macrophages Drive Tumor Inflammation in Uveal Melanoma. Mol Cancer Res, 2018. 16(12): p. 1902–1911.

30. Dietrich, D., et al., Improved PCR performance using template DNA from formalin-fixed and paraffin-embedded tissues by overcoming PCR inhibition. PLoS One, 2013. 8(10): p. e77771.

31. Kim, S.S., et al., Droplet digital PCR-based EGFR mutation detection with an internal quality control index to determine the quality of DNA. Sci Rep, 2018. 8(1): p. 543.

32. Lu, X.J.D., et al., Using ddPCR to assess the DNA yield of FFPE samples. Biomol Detect Quantif, 2018. 16: p. 5–11.

33. Jamal-Hanjani, M., et al., Translational implications of tumor heterogeneity. Clin Cancer Res, 2015. 21(6): p. 1258–66.

34. Shain, A.H., et al., Genomic and Transcriptomic Analysis Reveals Incremental Disruption of Key Signaling Pathways during Melanoma Evolution. Cancer Cell, 2018. 34(1): p. 45–55 e4.

35. Song, P., et al., Limitations and opportunities of technologies for the analysis of cell-free DNA in cancer diagnostics. Nat Biomed Eng, 2022. 6(3): p. 232–245.

36. Huggett, J.F., S. Cowen, and C.A. Foy, Considerations for digital PCR as an accurate molecular diagnostic tool. Clin Chem, 2015. 61(1): p. 79–88.

