## Supplementary Information for "Allele-specific digital PCR enhances precision and sensitivity in the detection and quantification of copy number alterations in heterogeneous DNA samples: an *in silico* and *in vitro* validation study"

### Supplementary Table 1

Context sequence, annealing temperature and available supplier information for digital PCR assays used in this study.

**Target assays**

| Assay | *PPARG* target chromosome 3p (FAM-labelled) |
|---|---|
| Context sequence | ATCAAAGTGG AGCCTGCATC TCCACCTTAT TATTCTGAGA AGACTCAGCT CTACAATAAG CCTCATGAAG AGCCTTCCAA CTCCCTCATG GCAATTGAAT GTCGTGTCTG TGGAGATAAA GCT |
| Location (hg19) | chr3:12422826-12422948 |
| Location (hg38) | chr3:12381327-12381449 |
| Amplicon length | 69 nucleotides |
| Annealing temperature | 60 °C |
| Supplier and assay ID | Bio-Rad (dHsaCP100462) |

| Assay | *PTK2* target chromosome 8q (FAM-labelled) |
|---|---|
| Context sequence | CAACCAGATG GTCATTCAAA AAAGTTGGAG CTGTAAGTGC TGGCGACTGA GGACACAGGG TTAATTCCTC GCTGCTGGTG GAAGGCTAGA GAACATCTTC AAAAGAGGGT AGCAAGACGT GCT |
| Location (hg19) | chr8:141669430-141669552 |
| Location (hg38) | chr8:140659331-140659453 |
| Amplicon length | 70 nucleotides |
| Annealing temperature | 60 °C |
| Supplier and assay ID | Bio-Rad (dHsaCP1000155) |

**Reference assays:**

| Assay | *TERT* reference chromosome 5p (HEX-labelled) |
|---|---|
| Context sequence | CACCCCTTGG TGGCGGCTCA CCTGTACGCC TGCAGCAGGA GGATCTTGTA GATGTTGGTG CACACCGTCT GGAGGCTGTT CACCTAGAGT CGCCAAGAAA GAGTGAGAAA CGGTAGAAAC CTC |
| Location (hg19) | chr5:1258692-1258814 |
| Location (hg38) | chr5:1258577-1258699 |
| Amplicon length | 100 nucleotides |
| Annealing temperature | 60 °C |
| Supplier and assay ID | Bio-Rad (dHsaCP1000100) |

| Assay | *VOPP1* reference chromosome 7p (FAM-labelled) |
|---|---|
| Context sequence | TATGGAGAGG GCCCGCACAC AGCACCTGGA GCCACAGCAG TCCTCGTAGG AGCGGCATCT GTGGAGAGAG GCACAGGCTG GTCAGCACTG AATTGGAAGC AGCCACCGGA CCAGCCATGC GGC |
| Location (hg19) | chr7:55565326-55565448 |
| Location (hg38) | chr7:55497633-55497755 |
| Amplicon length | 64 nucleotides |
| Annealing temperature | 60 °C |
| Supplier and assay ID | Bio-Rad (dHsaCP2506292) |

| | |
|---|---|
| Assay | *TTC5* reference chromosome 14q (HEX-labelled) |
| Context Sequence | TGGTCGCGAT GCCACTGTGG CAACAGCCTG GCTGCTGGAT CCCTGAGGCT TCCCATTCAC CACTAGCAGG AGGGGCGTCT CCACTCGAAC ACTGGAAAAG GAATAGTCCT AGAAAAGACA GAC |
| Location (hg19) | chr14:20757798-20757920 |
| Location (hg38) | chr14:20289639-20289761 |
| Amplicon length | 59 nucleotides |
| Annealing temperature | 60 °C |
| Supplier and assay ID | Bio-Rad (dHsaCP2506733) |

**SNP assays:**

| | |
|---|---|
| Assay | SNP rs2236947 (C/A) chromosome 3p (FAM/HEX-labelled) |
| Context sequence | CTCCTGGGCT AGGCACAAGA TCATTCTACA GGAAACCTTG TGGGAATTCT TCTGGGACAA A[G/T]TATTGGT CAGCGCTGAG CTTAGCTGTG TCTGTGACAC TCGCATTCTA ACTAGGGCCT ATCT |
| Location (hg19) | chr3:50371371-50371493 |
| Location (hg38) | chr3:50333940-50334062 |
| Amplicon length | 69 nucleotides |
| Annealing temperature | 55 °C |
| Supplier and assay ID | Bio-Rad (dHsaMDS958483376) |

| | |
|---|---|
| Assay | SNP rs1062633 (C/T) chromosome 3p (FAM/HEX-labelled) |
| Context Sequence | TGCTGGCAGC TGCACATAAT GGTCCCCAAG CAGTGCAGAC ACTATCTGCT CCACCTCCCC CACTAGTACT C[C/T]GAAGGTGG GTCGCACTGC TGGGTCTGCC TCCCAGCATT GCTGCATCAC TTGGTACCTG TTGGGGGAAA GGGATGTCAG GTTAAGGCAA TTTCCACCCA AGGATTCTGG GCCAC |
| Location (hg19) | chr3:49924869-49925053 |
| Location (hg38) | chr3:49887436-49887620 |
| Amplicon length | 130 nucleotides |
| Annealing temperature | 55 °C |
| Supplier | Sigma-Aldrich |

| | |
|---|---|
| Assay | SNP rs7018178 (C/T) chromosome 8q (FAM/HEX-labelled) |
| Context Sequence | CCATCGGCTC CTCCCGCCCA TGGGACCTTC GAAGACACTG CTGCCTCTGC CTGAAGTACT TTACCTTCCC CCCTTACCTG ACA[C/T]TCAATA ACTGCTTAGT GCCCTTAAAA GAGCTGCACT CTCAGGAGCA GAGTTTTGTG AGCATAGCAG GCTTCCAACT GACCCCTGGT TCCTCCCTCA |
| Location (hg19) | chr8:142239127-142239306 |
| Location (hg38) | chr8:141229028-141229207 |
| Amplicon length | 122 nucleotides |
| Annealing temperature | 55 °C |
| Supplier | Sigma-Aldrich |

| | |
|---|---|
| Assay | SNP rs7843014 (A/C) chromosome 8q (FAM/HEX-labelled) |
| Context Sequence | GGATCAGAGA GTGATGGGAC CTAAACCCAT TGATTAGTCT TAACATCACA AAGAACAACC AGATGTACAT CACCTA[A/C]GAA GTATTCTTGT TGTGGGGAGG GGGGGAATCA AGCCCAAACT GGATCAAACT TCTTTAGGTT |
| Location (hg19) | chr8:141790405-141790544 |
| Location (hg38) | chr8:140780306-140780445 |
| Amplicon length | 112 nucleotides |
| Annealing temperature | 55 °C |
| Supplier and assay ID | Sigma-Aldrich |

### Supplementary Figure 1

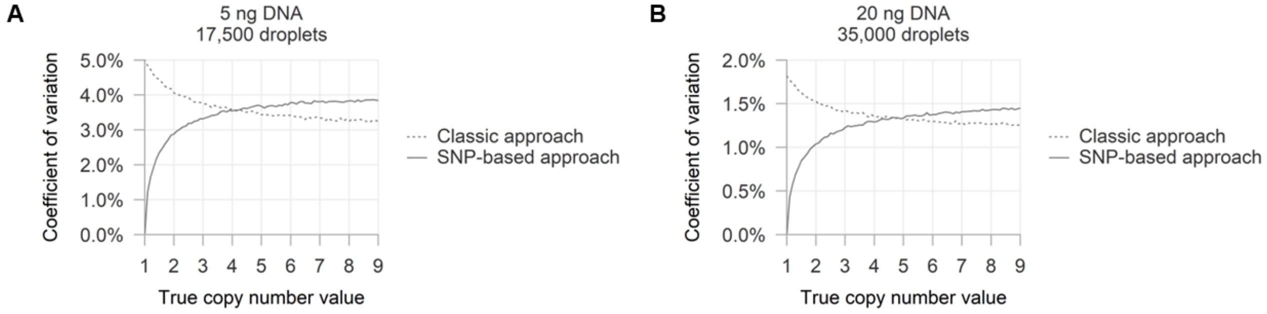


Performance of classic and SNP-based approaches in detecting and quantifying heterogeneous copy number alterations. All conditions are evaluated *in silico* using 10,000 simulations.

(**A**) Coefficient of variation of measured copy number values per simulated condition with 5 ng DNA and 17,500 droplets.

(**B**) Coefficient of variation of measured copy number values per simulated condition with 20 ng DNA and 35,000 droplets.

### Supplementary Data 1

#### Five phases of an *in vitro* digital PCR experiment

The workflow of an *in vitro* digital PCR experiment consists of five phases: (1) preparation of the reaction mixture, (2) partitioning of the reaction mixture (i.e. droplet generation), (3) PCR amplification, (4) fluorescence measurements (i.e. droplet reading) and (5) mathematical interpretation of the result (**Supplementary Data Figure 1**).

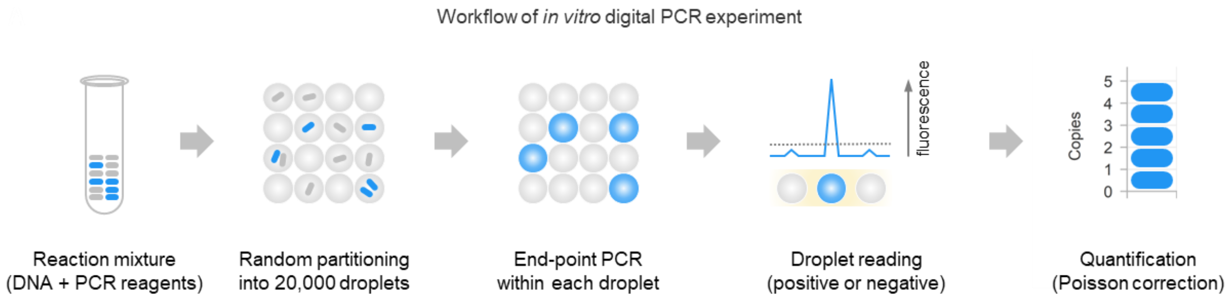


**Supplementary Data Figure 1** – Workflow of *in vitro* digital PCR experiment.

The reaction mixture contains of a DNA sample (e.g. 20 ng genomic DNA), primers and probe(s) for one or more DNA targets of interest, and other required PCR reagents. Next, this mixture is distributed randomly over a large number of partitions, which are ~20,000 droplets when using the Droplet Digital PCR system (Bio-Rad). As typically the total number of droplets is larger than the number of DNA target molecules, only a fraction of the droplets contains a DNA target of interest. During the PCR, amplification only takes place in those droplets and results in a high fluorescence intensity (droplets scored as ‘positive’), which is measured in the reading phase. In contrast, droplets without initial presence of a DNA target molecule show a low background level of fluorescence and are scored as ‘negative’ for the chosen DNA target.

The number of positive droplets reflects the abundance of the measured DNA target: the more target molecules are to be distributed, the more droplets will be filled and eventually scored as positive. However, this relationship is not linear, as the random DNA distribution can also lead to droplets containing more than one target molecule (**Supplementary Data Figure 2**).

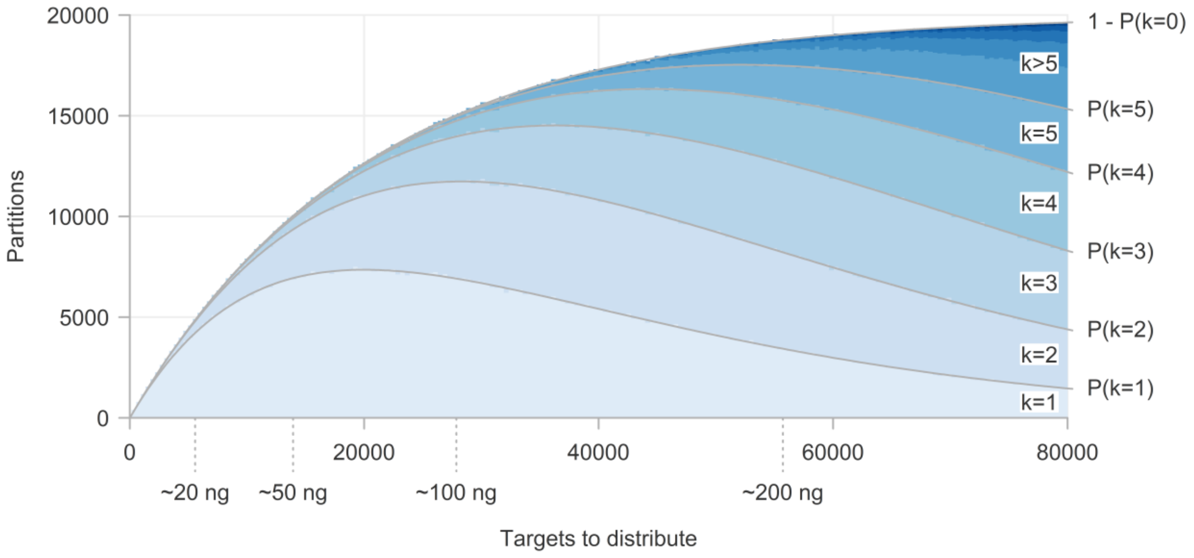


**Supplementary Data Figure 2** – Average distribution of $x$ targets over 20,000 partitions.

Instead, the relationship between positive droplets and number of target molecules follows a Poisson distribution [8, 12, 13]. Because an end-point PCR is performed, all positive droplets have a comparable fluorescence intensity, independently of the initial number of DNA target molecules present. 'Digital' in digital PCR refers to this dichotomous way of scoring: each droplet can only be positive or negative for a certain target. For this reason, the final phase of a digital PCR experiment involves a careful mathematical interpretation of the obtained results.

**Mathematical interpretation of a digital PCR experiment**

The probability of finding a number of (initial) targets per droplet ($\kappa$) can be described following the Poisson distribution:

$$P(\kappa) = \frac{\lambda^k}{\kappa!} e^{-\lambda}$$

…where $e$ is Euler's number ($e = 2.71828...$) and $\lambda$ is the expected (i.e. average) number of targets per droplet.

In **Supplementary Data Figure 2**, a simulated number of targets ($n$, on the $x$-axis) is randomly distributed over 20,000 virtual droplets, which allows us to determine the average number of targets per droplet:

$$\lambda = \frac{n}{20{,}000}$$

P($\kappa$) can now be calculated for all values of *n* and increasing values of $\kappa$ (lines in **Supplementary Data Figure 2**). The high concordance between these calculated and observed values confirms that the Poisson distribution can be used to estimate the number of targets per droplet.

In a digital PCR experiment, one is particularly interested to estimate $\lambda$, as its value can be used to calculate the true number of DNA target molecules (*n*). However, only the observed fraction filled $P(\kappa \geq 1)$ and fraction empty droplets $P(\kappa = 0)$ are known. These values can still be used to estimate $\lambda$:

$$\begin{aligned} P(\kappa = 0) &= \frac{\lambda^0}{0!} e^{-\lambda} = e^{-\lambda} \\ \log\left(P(\kappa = 0)\right) &= -\lambda \\ \lambda &= -\log\left(P(\kappa = 0)\right) \\ \lambda &\approx -\log\left(1 - \frac{\text{number of droplets target positive}}{\text{total number of droplets}}\right) \end{aligned}$$

**Performing *in silico* digital PCR experiments**

To perform digital PCR experiments *in silico* (**Figure 1A**), we developed R library *digitalPCRsimulations*, which is available via https://github.com/rjnell/digitalPCRsimulations:

```
# Install library
library(devtools)
install_github("rjnell/digitalPCRsimulations")

# Load library
library(digitalPCRsimulations)

# Set random seed
set.seed(123)
```

To simulate a typical singleplex digital PCR experiment, virtual universe $u_1$ (which has an arbitrarily large size of 100x 20,000 droplets) can be created and filled with *n* targets per 20,000 droplets, where *n* is equivalent to the number of genomic copies in a certain amount of input DNA (e.g. 20 ng), based on the weight of one healthy haploid genome ($3.59 \cdot 10^{-3}$ ng).

```r
# Function universe(...)
## input: amount of input DNA (in nanograms)
## [optional] n_size: size of universe (x 20,000 droplets)
u_1 = universe(input = 20,
               n_size = 100)
```

Next, a random sample of *m* droplets can be taken from universe $u_1$ and the number of positive droplets (i.e. containing one or more targets) is measured and interpreted mathematically in a similar way as the result of an *in vitro* digital PCR experiment. With only a fraction of the universe of droplets being measured, subsampling of the DNA sample is mimicked *in silico*. Moreover, as *in vitro* experiments usually yield less than 20,000 droplets (i.e. part of the droplets is not measured successfully and left out from downstream analyses), *m* can be set to a more realistic value of 17,500 droplets as desired:

```r
# Number of droplets to analyse
m = 17500

# Function sample_from_universe(...)
## universe: an existing universe
## sample: size of the sample to be taken
## [optional] alpha: level of significance of returned confidence interval
concentration = sample_from_universe(universe = u_1,
                                     sample = m,
                                     alpha = 0.05)
```

Based on the number of positive (and total) droplets from the random sample, the concentration of target molecules (in copies/uL) is automatically estimated using the Poisson distribution and the known volume of one droplet (0.85 nL, Bio-Rad). Depending on the level of significance, a confidence interval is calculated as well (with *alpha* = 0.05, a 95%-confidence interval is calculated):

```r
concentration
#>   concentration   concentration_low   concentration_high
#>        329.9648            320.0885             339.9246
```

Additionally, duplex digital PCR experiments, measuring two independent targets, can be simulated by creating two universes and taking an equally-sized sample of both:

```r
# E.g. two targets that are equally abundant ~ number of genomic copies in 20 ng:
u_duplex_1 = universe(input = 20)
u_duplex_2 = universe(input = 20)
# Estimation of concentrations of individual targets
concentration_duplex_1 = sample_from_universe(universe = u_duplex_1,
                                              sample = m,
                                              alpha = 0.05)
concentration_duplex_2 = sample_from_universe(universe = u_duplex_2,
                                              sample = m,
                                              alpha = 0.05)
```

Note that, while both targets are equally abundant, random subsampling and partitioning results in different individual concentrations:

```
concentration_duplex_1
#>   concentration   concentration_low   concentration_high
#>        329.0752            319.2141             339.0196

concentration_duplex_2
#>   concentration   concentration_low   concentration_high
#>        323.1323            313.3732             332.9730
```

The difference between both concentrations can be measured by calculating a ratio of both with its confidence interval (following Fieller's theorem):

```
# Function calc_ratio(...)
## input_a: numerator concentration and lower/upper confidence interval limit
## input_b: denominator concentration and lower/upper confidence interval limit
ratio = calc_ratio(input_a = concentration_duplex_1,
                   input_b = concentration_duplex_2)
```

As in this case value 1 is within the limits of the confidence interval, the difference between both targets is not statistically significant:

```
ratio
#>       ratio   ratio_low   ratio_high
#>   1.0183915   0.9757958    1.0628608
```

The previous steps can be combined with the following shortcut function:

```
# Function simulate_ratios(...)
## universe_1: an existing universe
## universe_2: an existing universe
## n_droplets: size of the sample to be taken
## n_simulations: number of simulations
## [optional] alpha: level of significance of returned confidence interval
duplex_ratios = simulate_ratios(universe_1 = u_duplex_1,
                                universe_2 = u_duplex_2,
                                n_droplets = m,
                                n_simulations = 50,
                                alpha = 0.05)
```

The resulting ratios with associated confidence intervals are saved in a matrix and can be visualised in a plot. Note that value 1 is not within the limits of all confidence intervals:

```
duplex_ratios[1:10,]
#>                      ratio   ratio_low   ratio_high
#>   simulation-1    1.0181020   0.9755353     1.062540
#>   simulation-2    1.0000000   0.9580896     1.043744
#>   simulation-3    1.0237440   0.9814163     1.067915
#>   simulation-4    1.0087724   0.9664923     1.052909
#>   simulation-5    1.0176837   0.9753290     1.061891
```

```
#>   simulation-6    0.9527998   0.9128222     0.994493
#>   simulation-7    0.9970195   0.9553684     1.040484
#>   simulation-8    1.0021696   0.9603456     1.045817
#>   simulation-9    1.0222199   0.9795279     1.066790
#>   simulation-10   1.0100653   0.9679356     1.054036

# Function plot_simulations(...)
## results: numeric matrix with individual results and confidence intervals
## [optional] main: title of plot
## [optional] xlab: label of x-axis
## [optional] ylab: label of y-axis
## [optional] ylim: vector with lower and upper limit of y-axis
## [optional] true_value: true value to compare results to
## [optional] error: logical to plot confidence intervals
## [optional] reverse: logical to reverse default colour palette
## [optional] sort: logical to sort obtained results before plotting
plot_simulations(results = duplex_ratios,
                 main = "Results",
                 xlab = "Simulation",
                 ylab = "Ratio",
                 ylim = c(0.9,1.1),
                 true_value = 1,
                 error = T,
                 reverse = F,
                 sort = T)
```

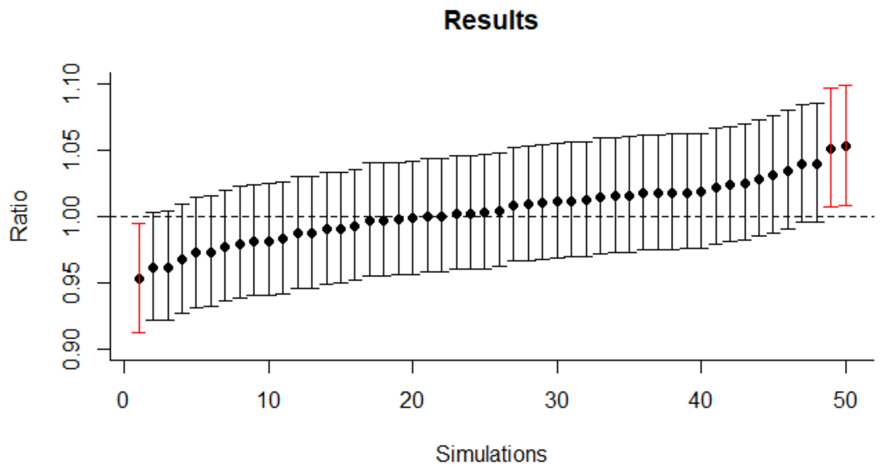


**Supplementary Data Figure 3 –** Simulation results of experiments measuring a ratio of 1.

A variety of statistics can be calculated on the results of the simulated experiments:

- number of experiments with a confidence interval not containing the true value;
- number of experiment with a confidence interval containing the true value;
- fraction experiments with a confidence interval containing the true value;
- point estimate of the mean of the results;
- point estimate of the standard deviation of the results.

In this example, the point estimate mean of the ratios is close to our true value of 1. Moreover, the confidence intervals contained the true value of 1 in 94% the experiments, which is close to the intended 95%:

```
# Function stats(...)
## results: numeric matrix with individual results and confidence intervals
## true_value: true value to compare results to
stats(duplex_ratios, true_value = 1)
#>   $not_in_interval
#>   [1] 3
#>
#>   $in_interval
#>   [1] 47
#>
#>   $coverage
#>   [1] 0.94
#>
#>   $point_estimate_mean
#>   [1] 1.003648
#>
#>   $point_estimate_sd
#>   [1] 0.0228502
```

Finally, the described functions can be used to model the classic and SNP-based approach and simulate the digital PCR analysis to determine copy number values:

```
# INPUT VALUES
# What is the true copy number value to be simulated?
cnv = 1.9

# What is the DNA input (in ng)
input_ng = 20

# What is the number of droplets per experiment?
n_droplets = 17500

# How many simulations per condition should be performed?
n_simulations = 50

# Which alpha should be used?
alpha = 0.01
```

```
# CLASSIC APPROACH
# Create universes
universe_cnv = universe(input_ng * cnv/2)
universe_ref = universe(input_ng)

# Simulate classic CNV experiment
classic_cnv = simulate_ratios(universe_cnv,
                              universe_ref,
                              n_droplets = n_droplets,
                              n_simulations = n_simulations,
                              alpha = alpha)
```

```r
# Transform ratio to CNV
classic_cnv = cbind(classic_cnv[, 1]*2,
                    classic_cnv[, 2]*2,
                    classic_cnv[, 3]*2)

# Calculate simulation statistics
classic_cnv_stats = stats(classic_cnv, true_value = cnv)

# In which % would we classify the result as not-normal?
classic_cnv_ci_sensitivity = 1-stats(classic_cnv, true_value = 2)$coverage
```

```r
# SNP-BASED APPROACH
# Calculate number of copies target per haploid genome equivalent
cnv_snp1 = max(0, cnv-1)
cnv_snp2 = 1

# Create universes
universe_snp1 = universe(input_ng * cnv_snp1/2)
universe_snp2 = universe(input_ng * cnv_snp2/2)

# Simulate new SNP experiment
if (cnv-1 < 0) {
  snp_cnv = cbind(rep(c(NA, NA, NA), n_simulations))
} else {
  snp_cnv = simulate_ratios(universe_snp1,
                            universe_snp2,
                            n_droplets = n_droplets,
                            n_simulations = n_simulations,
                            alpha = alpha)
  snp_cnv = cbind(snp_cnv[, 1]+1,
                  snp_cnv[, 2]+1,
                  snp_cnv[, 3]+1)
}

# Calculate simulation statistics
snp_cnv_stats = stats(snp_cnv, true_value = cnv)

# In which % would we classify the result as not-normal?
snp_cnv_ci_sensitivity = 1-stats(snp_cnv, true_value = 2)$coverage
```

The results of these simulations show a notable difference in sensitivity between the classic and SNP-based approach, as further illustrated, validated and discussed in the main text of this manuscript:

```r
# COMPARISON OF RESULTS
# Sensitivity of classic approach
classic_cnv_ci_sensitivity
#> [1] 0.5

# Sensitivity of SNP-based approach
snp_cnv_ci_sensitivity
#> [1] 0.88
```

```
# Plot of classic approach
plot_simulations(classic_cnv,
                 main = "Classic approach",
                 xlab = "Simulations",
                 ylab = "Copy number value",
                 ylim = c(1.7,2.2),
                 true_value = 2,
                 error = T,
                 reverse = T,
                 sort = T)

# Plot of SNP-based approach
plot_simulations(snp_cnv,
                 main = "SNP-based approach",
                 xlab = "Simulations",
                 ylab = "Copy number value",
                 ylim = c(1.7,2.2),
                 true_value = 2,
                 error = T,
                 reverse = T,
                 sort = T)
```

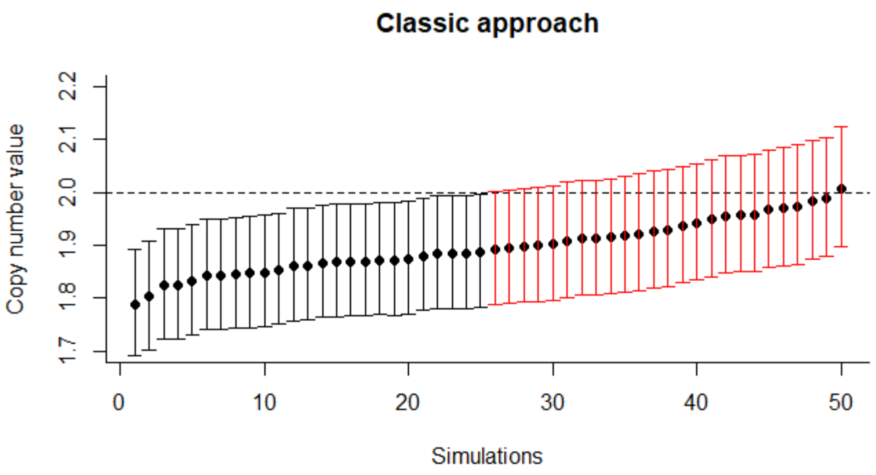


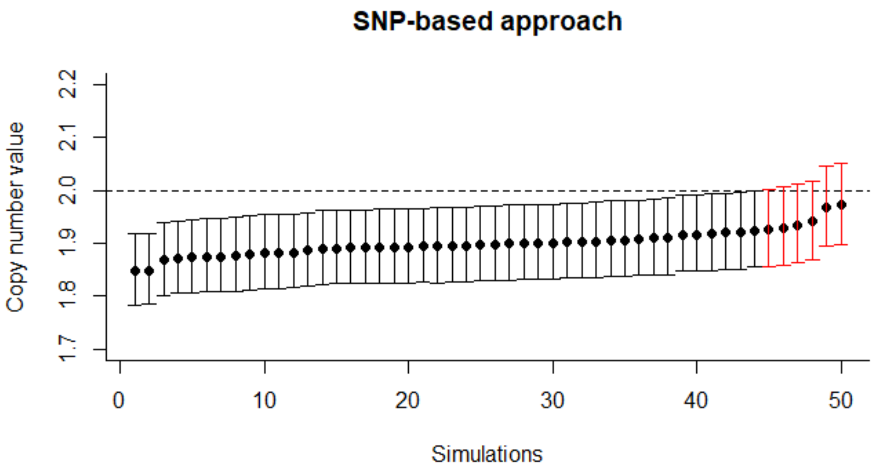


**Supplementary Data Figure 4 –** Comparison of simulation results of classic and SNP-based approach to determine a copy number value of 1.9.
